# Internet Delivered Sexually Transmitted Infection and Teen Pregnancy Prevention Program: A Randomized Trial

**DOI:** 10.1101/2022.09.13.22279867

**Authors:** Patricia J. Kissinger, Jakevia Green, Jennifer Latimer, Norine Schmidt, Aneeka Ratnayake, Aubrey Spriggs Madkour, Gretchen Clum, Gina M Wingood, Ralph J. DiClemente, Carolyn Johnson

## Abstract

**Importance:** Black teenaged women have disproportionately high rates of unintended pregnancy (UTP) and sexually transmitted infections (STI). Internet-based interventions can be delivered to large groups of people in a relatively inexpensive manner.

**Objective:** The purpose of this randomized trial was to examine the efficacy of an adaptation of a theory- and evidence-based UTP/STI prevention intervention for older teens and for internet delivery.

**Design:** A randomized, attention-controlled trial.

**Setting:** Participants were recruited from minority-serving institutions and community events in New Orleans, Louisiana.

**Participants:** Black women aged 18-19 who were not pregnant nor seeking to become pregnant were enrolled (n=637).

**Main outcomes and measures:** Participants were randomized to an 8-session intervention or attention control and were followed at 6- and 12-months post-intervention to determine if the use of reliable contraception, dual methods, intention to use condoms, and intention to use reliable contraception were higher among women in the intervention arm compared to controls. Pregnancy and STI rates were also compared.

**Results:** Overall, at baseline, reliable contraception was 54.8% and dual protection was 29.4% and the prevalence of STI was 11.1%. Participants were similar by arm. Participation and follow-up rates were excellent (60.9% and 80.3%). At 6 months, all outcomes were improved in intervention compared to control, but this effect waned by 12 months.

**Conclusion and Relevance:** The intervention was efficacious for increasing self-reported pregnancy and STI prevention behavior and intentions and lower trends for pregnancy and STIs were observed in this group which decreased over time suggesting the need for a booster.

**Trial Registration:** ClinicalTrial.gov NCT01579617

## Introduction

While teen birth rates fell to an historic rate in 2017, they remain higher than most western countries^1^ and are likely to rise with the reversal of federal protection for abortion rights by the Unites States Supreme Court in 2022.^2^ Teen births are highest in the older teen ages with those 18-19 having a three-fold higher pregnancy rate than teens 15-17.^3^ Teen pregnancy disproportionately affects youth of color; young teens who are black become pregnant 2.6 times more often than those who are white.^4^

UTPs and STIs have individual and societal consequences. Children of teen mothers are more likely to live in poverty, have lower academic achievement, and experience an UTP themselves than children born to non-teen mothers.^5^ Women with chlamydia or gonorrhea infections are more likely to develop pelvic inflammatory disease, experience ectopic pregnancy, become infertile, and acquire HIV than those without these infections.^6^ The financial burden of these health conditions is enormous. The annual cost of UTP and STIs among youth to U.S. taxpayers has been estimated to be 9.4 billion dollars.

Louisiana is one of 19 states that does not mandate sex education or HIV prevention education in schools, not does it allow for the distribution of condoms or contraception in schools.^7^ Louisiana consistently ranks high for sexually transmitted infections and teen births. In 2020 among all states, Louisiana ranked 2^nd^ for chlamydia and gonorrhea, and 3^rd^ highest rate of teen births.^8, 9^

Many older teenaged women have not received essential knowledge and skills to protect themselves against UTP and STIs. Half of UTPs occur among teenaged women who use contraception inconsistently or incorrectly,^10^ suggesting an ongoing need for teens to obtain factual education and prevention skills regarding sexual intercourse and contraception. Interventions to prevent UTP, HIV, and STIs are greatly needed, particularly in states where sex education is suboptimal.

The purpose of this study is to evaluate the efficacy of a theory-based internet-delivered pregnancy and STI intervention for older teenaged Black women using a randomized controlled study design and biological outcomes.

## Methods

### Intervention Description

Be yoU, Talented, Informed, Fearless, Uncompromised, and Loved (BUtiful) is an internet-delivered pregnancy prevention intervention. BUtiful was an adaptation of the *Sisters, Informing, Healing, Living, and Empowering* (SiHLE) that was found to reduce STIs and unintended pregnancies.^11^ SiHLE is a 4 session face-to-face intervention for HIV and STI prevention among Black adolescent females aged 14-18 and is based on the Social Cognitive Theory and the Theory of Gender and Power.^12^ To create the BUtiful intervention, three adaptations to SiHLE were made using the ADAPT-IT methodology and in collaboration with the creators of SiHLE.^13^ These adaptations were: targeting older teenage women aged 18-19, including more information on contraception, and changing from face-to-face group session delivery to asynchronous internet delivery via a website. It was designed to be conveniently disseminated to older teenaged women in various settings and would assure standardization of the content, provide flexibility of delivery and be appealing to youthful audiences.

The BUtiful website was composed of eight successive interactive sessions that participants could access at their convenience within four weeks (Table 1). Participants’ access to the website was activated on the day of enrollment and was deactivated at the conclusion of four weeks. Each session took an average of 30 minutes to complete, depending on the participant’s level of interaction. The sessions were presented via video, text, interactive activities, and message boards. Five female characters presented the material in the sessions: four prototypic older teenage women of the target demographic who presented their experiences and worked through issues related to topics such as contraception and relationships, and one slightly older moderator whose character presents medically-accurate health information and anecdotes of her experiences with UTP and an STI.

**Table 1.** Description of intervention sessions

| Lesson Title | Lesson Description |
| --- | --- |
| BUtiful Beginnings | Introduction to website and characters, role models, women in music and media |
| Be yoU! | Understanding values and setting goals |
| BUtiful Body | Female reproductive anatomy, menstrual cycle |
| BUtiful Choices | Hormonal, non-hormonal, barrier, and natural birth control options and availability |
| BUtiful and Informed | STI/HIV information and prevention |
| BUtifully Communicate | Types of communication and negotiation strategies |
| BUtiful Relationships | Relationship choices, abuse and substance use |
| We are BUtiful | Wrap-up/Teach-back |

Study staff were available to answer questions about the site and to manage technical difficulties such as lost passwords. The website was password-protected and participants were required to sign a contract that they would not share the website with others. Study staff maintained contact with the participants throughout the intervention period through text messages, telephone calls, and email according to participant preference, to remind them of the four-week timeline and verify or update their contact information. Following the four weeks of site access, session material was reinforced in quarterly newsletters of which participants could opt out. This outreach was provided throughout the study (12 months post intervention). No modifications of the intervention were made during implementation.

### Attention control

The attention control arm created by the study team was composed of eight sequential interactive sessions that contained content distinct from BUtiful, instead delivering a general health and nutrition curriculum. Dosage and delivery mirrored the intervention methods, as did the types of activities and format. Four prototypic female characters and one older moderator were also followed using video. The technical assistance in accessing the website, and the reinforcement of information through quarterly newsletters after the four-week period were similar to the intervention arm.

### Study Design

The evaluation was a parallel randomized controlled trial. Sealed randomization cards, using block randomization generated by SAS statistical software, were prepared prior to the start of enrollment by staff who were not involved in recruitment

### Inclusion/exclusion

Women were eligible if they met the following criteria: identified as Black or African American, were 18 or 19 years of age at the time of enrollment, were not pregnant at recruitment (per urine pregnancy test), did not intend to become pregnant within a year’s time, were living in Orleans or Jefferson Parish (County), were not previously enrolled, did not exclusively have sex with women, and did not have a female roommate or relative participating in the program (to avoid the risk of contamination).

### Recruitment

Subjects were recruited from 9/7/12 to 9/30/14 from diverse locations including a community college with two campuses, three historically Black universities and one adolescent drop-in health center. Women were also recruited from community settings at multiple venues, such as beauty shops and community events.

### Enrollment

Enrollment was conducted using a phase-in approach. Women who were interested and signed up for the study were either immediately enrolled or contacted within 2 to 3 days (with a maximum of 7 days) by study staff. Staff reviewed eligibility criteria and described the study to each interested woman. If a woman was interested and eligible, an enrollment visit was scheduled at a location convenient to her to obtain written informed consent. During the consenting process at the enrollment visit, women were told that the purpose of the study was to evaluate two online health programs for Black women that were similar in structure and time commitment but contained different health content, and that there was a 50/50 chance of being randomized to one online program or the other.

### Surveys

After obtaining informed consent, a baseline survey was administered using audio computer-assisted self-interview (ACASI). The survey elicited information on demographics, sexual history, coercive sexual experiences, substance use, sexual behaviors at the level of the male partner (up to four), contraceptive use and adherence, general health information, diet, and exercise. At baseline, a battery of questions and indices were also asked including: pregnancy and condom-use intentions, Positive Orientation for Early Motherhood scale (POEM),^14^ pregnancy prevention knowledge which was developed by the investigators, contraceptive self-efficacy,^15^ social desirability,^16^ depression using the CESD-10,^17^ perceived social support,^18^ self-esteem,^19^ substance use.^20^

### Biological testing

Participants provided a urine specimen that was collected to assess pregnancy, using a commercially available early pregnancy test for human chorionic gonadotropin (hCG), and tested for *Chlamydia trachomatis* (CT) and *Neisseria gonorrhoeae* (GC), using nucleic acid amplification testing (NAAT). Women who tested positive for pregnancy or an STI were referred to one of several clinics that provided low or no cost care (for those without insurance) or encouraged to seek follow-up care with their regular medical provider, depending on their preference. Women who tested positive for pregnancy via the study test were not eligible to continue in the study.

Following completion of baseline assessment procedures, study staff opened a sequentially numbered and sealed randomization envelope to reveal the participant’s assignment to study arm. Staff then demonstrated the assigned website and helped with log on procedures, collected contact information, and set up the three follow-up visits.

### Follow-up visits

Website activity was recorded during the four weeks of program delivery using Google Analytics™. Women were followed at 2-months (check in visit), 6-months (short--term outcomes), and 12-month (long-term outcomes) post-intervention and were re-interviewed via ACASI and retested for pregnancy and STIs. If a woman missed a follow-up visit, she was still pursued at the next visit. Satisfaction with the intervention was ascertained via survey. If participants did not complete the 2-month visit, relevant questions were moved to the next survey. Women who relocated from the area or who could not attend a visit were offered the option of completing their ACASI via the internet and sent a specimen collection kit via FedEx.

### Outcomes

Four primary outcomes were considered at 6 and 12 months. These included: 1) initiating reliable contraception use (which included any hormonal method or consistent condom use with all partners or sexual abstinence), 2) initiating dual method (reliable contraception plus condom use for every sex act), 3) intention to use condoms, and 4) intention to use reliable contraception in the next year.

There were two secondary outcomes: the presence of an STI (via Ct or GC NAAT testing or self-report of treatment for an interim Ct or GC infection during follow-up) and pregnancy (per pregnancy test or self-report of interim pregnancy).

### Sample size

The sample size was calculated assuming a 40-60% prevalence of the primary outcomes at baseline and a 35% increase as a result of the intervention. At an alpha of 0.05 for power of 0.80 accounting for a 20% or less loss-to-follow-up, the number of evaluable units needed in each arm was between 98 to 230.

### Data analysis

Data for the main outcomes were analyzed using intent-to-treat analysis. For both analyses, missing outcome data were imputed using Proc MI in SAS using 5 imputations per outcome. Generalized estimating equations was then performed to determine the effect measures, confidence intervals, and p-values. Because pregnancy could span several visits, Cox regression time-to-event analysis was conducted to evaluate the intervention effects on pregnancy.

### Human Subjects and study compensation

For their participation in the study, women were compensated $25 for the baseline visit, $50 for the 2-month, $75 for both the 6- and 12-month visits. Additionally, as incentive to complete the program, women in both arms were offered $5 for each of the eight sessions they completed. Session completion was tracked using hit counters on the last page of each session and monitored by study staff. Reminders to complete sessions and session completion tallies (including dollars earned) were sent to the participants via their preferred method of contact. This study was approved by the Tulane University Institutional Review Board 11-261904; registered in ClinicalTrial.gov NCT01579617; and a Certificate of Confidentiality was obtained from the U.S. Department of Health and Human Services (CC-HD-15-072).

## Results

### Demographics

There were 656 women who provided consent and were randomized. Of those, 19 were found to be ineligible post randomization including 3 who were found to be pregnant via urine test, 2 who were found to have sex with women exclusively, 1 who was out of the age range and another 13 who had an invalid consent or HIPAA form. Of the remaining 637, 315 were randomized to intervention arm and 322 to control arm. (Figure 1).

**Figure 1:**
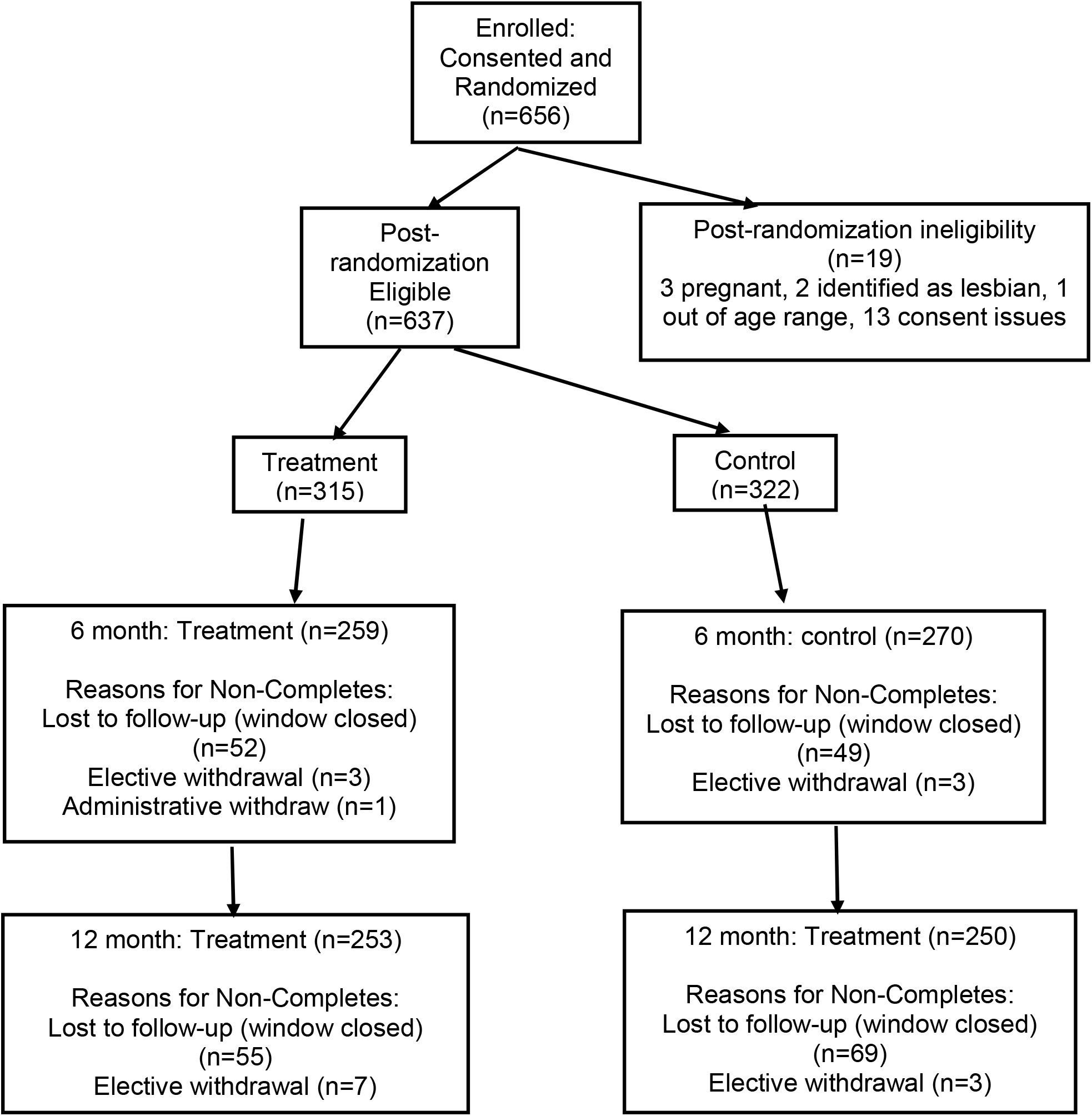
Consort Table

At baseline, the women were similar by demographics, sexual behavior, potentially confounding variables, STIs, and contraception use (Table 1). They were also similar by the battery of indices that could potentially influence the outcomes of interest (Table 2).

**Table 2.** Comparison of selected factors at baseline by arm.

|  | Intervention<br>(n=315)<br>% | Control<br>(n=322)<br>% | Total<br>(n=637)<br>% | p-value |
| --- | --- | --- | --- | --- |
| ***Sociodemographic Characteristics*** |  |  |  |  |
| 18 years old (versus 19) | 50.5 | 57.8 | 54.2 | 0.07 |
| Only has sex with men | 81.6 | 85.1 | 83.4 | 0.24 |
| Employed | 41.3 | 33.6 | 37.4 | 0.05 |
| In school | 98.1 | 97.2 | 97.6 | 0.46 |
| Completed high school/GED | 92.7 | 92.2 | 92.5 | 0.83 |
| Mother's education > high school | 59.5 | 64.6 | 62.1 | 0.19 |
| Father's education > high school | 41.9 | 48.7 | 45.4 | 0.09 |
| Live with family /relative | 60.5 | 66.9 | 63.7 | 0.10 |
| ***Sexual behaviors*** |  |  |  |  |
| Ever had vaginal sex | 75.0 | 76.7 | 75.9 | 0.62 |
| Ever anal sex | 10.2 | 8.7 | 9.4 | 0.51 |
| Ever oral sex | 55.8 | 53.7 | 54.7 | 0.61 |
| Age at sexual debut < 16 years of age | 22.0 | 26.4 | 24.2 | 0.19 |
| Ever pregnant | 7.7 | 11.2 | 9.4 | 0.13 |
| Ever gave birth | 4.8 | 5.3 | 5.1 | 0.79 |
| Ever diagnosed with CT or GC | 14.0 | 15.5 | 14.8 |  |
| Multiple sex partner in last three months | 54.6 | 45.7 | 50.1 | 0.02 |
| Definitely/Probably will have vaginal sex with man in next 12 months | 69.7 | 70.8 | 70.3 | 0.77 |
| Trying to avoid pregnancy | 94.9 | 94.4 | 94.7 | 0.77 |
| ***Potentially confounding covariates*** |  |  |  |  |
| Friend had child before age 20 | 30.5 | 33.9 | 32.2 | 0.36 |
| Sister had child before age 20 | 19.0 | 19.6 | 19.3 | 0.87 |
| Mom had child before age 20 | 48.9 | 44.1 | 46.5 | 0.23 |
| Person who referred was in the study | 7.1 | 11.0 | 9.1 |  |
| Depression per CESD-10 | 47.2 | 43.7 | 45.5 | 0.37 |
| Binge drinking in last 3 months | 26.7 | 23.9 | 25.3 | 0.42 |
| Used marijuana in past 3 months | 43.3 | 47.7 | 45.5 | 0.27 |
| Was in prior HIV/STI program | 9.2 | 8.4 | 8.8 | 0.72 |
| **Baseline STDs** |  |  |  |  |
| *Chlamydia trachomatis* | 10.2 | 9.9 | 10.0 | 0.93 |
| *N. gonorrhoeae* | 1.3 | 1.6 | 1.4 | 0.76 |
| **Baseline contraceptive use** |  |  |  |  |
| Long acting reliable | 3.5 | 5.3 | 4.4 | 0.27 |
| Other contraceptive | 47.6 | 51.9 | 49.8 | 0.28 |
| Consistent condom use or abstinence | 75.6 | 73.9 | 75.2 | 0.45 |
| Dual protection | 35.6 | 29.8 | 29.4 | 0.84 |

Per inclusion criteria, all women were Black/African American, with 3.6% also identifying with a second race/ethnicity (1.7% Hispanic, 1.3% American Indian, 0.2% Asian, 1.1% other non-specified). Over half (54.2%) were 18 years compared to 19 years old. The majority were in school (97.6%), most had completed high school or equivalent (98.8%), and 37.4% were employed. Only 18 of the women worked full time (all of whom were in the control arm).

### Sexual history

The majority of women previously engaged in a form of sex, including vaginal (75.9%), oral (54.7%), and anal (9.4%), and 24.2% had their sexual debut before the age of 16 (the average age of sexual debut for a Black women in the US).^21^ Some (9.4%) reported a history of pregnancy, 5.1% had given birth, and 14.8% reported a history of Ct or GC. All denied having ever tested positive for HIV. About half (50.1%) had multiple sexual partners in the 3 months prior to enrollment, and 70.3% reported that they definitely or probably would have sex with a man in the next 12 months. While, per protocol, all women were not trying to get pregnant, 5.4% of the said they would not mind getting pregnant if it happened.

### Potential confounders

When asked about female friends and relatives that had a child before the age of 20, women responded “yes” to a friend (67.8%), a sister (19.3%), or their mother (46.5%). Nearly half (45.5%) had a high score of depressive symptoms per the CESD-10 (a score of 10 or higher), and 25.3% said they binge drank in the last 30 days.

The overall mean for POEM scale was low, 16.5 (range 8-40) indicating disinterest in becoming a mother at this time; pregnancy prevention knowledge was high, 6.2 (range 0-9); and contraceptive self-efficacy was high, 72.8 (range 18-90), as was self-esteem, 33.1 (range 14-40). Social desirability fell in the middle, 5.6 (range 0-10), as did social support, 65.6 (range 12-98). These scores did not differ by arm.

### Outcomes at baseline

None of the women included in the analysis were pregnant at baseline, per protocol; however, 10.2% tested positive for CT and 1.4% with GC. Self-reported use of reliable contraceptives was 54.8%, consistent condom use was 72.4%, and dual protection was 29.4%.

### Missed visits

Of the 637 women followed, 76.1% attended all follow-up visits. Compared to those who attended all visits, those who missed visits tended to be less likely to have a job (31.6% vs. 39.3%, p=0.09), were less likely to be a high school graduate (88.2% vs.93.8%, p=0.02), and tended to be less likely to have multiple partners (44.1% vs. 52.0%, p=0.09). Those who missed a visit were similar to those who did not miss a visit on all other variables presented in Tables 2-3 at the p>0.10 level. Intervention women were also as likely as control women to attend their 6-month visit (82.2% vs. 83.9%, p=0.82) and 12-month visit (80.6% vs. 77.6%, p=0.29).

**Table 3.** Scales at baseline by arm.

|  | Range | Intervention<br>(n=315)<br>Mean (S.D.) | Control<br>(n=322)<br>Mean (S.D.) | Total<br>(n=637)<br>Mean (S.D.) | p-value |
| --- | --- | --- | --- | --- | --- |
| Positive Orientation for Early Motherhood | 8-40 | 16.5 (7.1) | 16.7 (7.8) | 16.6 (7.5) | 0.77 |
| Pregnancy Prevention Knowledge | 0-9 | 6.2 (1.5) | 6.3 (1.4) | 6.2 (1.5) | 0.57 |
| Contraceptive Self-Efficacy | 18-90 | 72.6 (8.7) | 72.3 (9.5) | 72.4 (9.1) | 0.66 |
| Social Desirability | 0-10 | 5.4 (2.2) | 5.7 (2.1) | 5.6 (2.1) | 0.05 |
| Social Support | 12-98 | 65.9 (15.4) | 65.3 (16.5) | 65.6 (15.9) | 0.65 |
| Self Esteem | 14-40 | 33.2 (4.8) | 33.1 (5.3) | 33.1 (5.1) | 0.88 |

### Adherence to program

Of 637 women in the study, 23.7% did not engage in any sessions, while 60.7% engaged in 80% or more of the sessions. Women in the intervention arm were as likely as those in the attention-control to complete 80% or more of the sessions (58.1% vs. 63.7%, p<0.15). Most participants (78.2%) opted to receive the newsletter, but there was no difference by arm in opt-in rate (79.4% in the intervention vs. 71.0% in the control, p=0.47). There was also no mechanism to know if they read the newsletter.

### Primary outcomes

In the intent-to-treat analysis, at 6 months, women in the intervention arm were more likely than those in the comparison group to start using reliable contraception and to start using dual methods (p<0.06). Intervention women were also more likely to intend to use condoms and reliable contraception in the next year (p< 0.03). While this trend continued at 12 months, only intention to use condoms remained statistically significant (Table 4 and Figure 2). A similar pattern was found for the per protocol analysis (Table 4 and Figure 3).

**Table 4.**
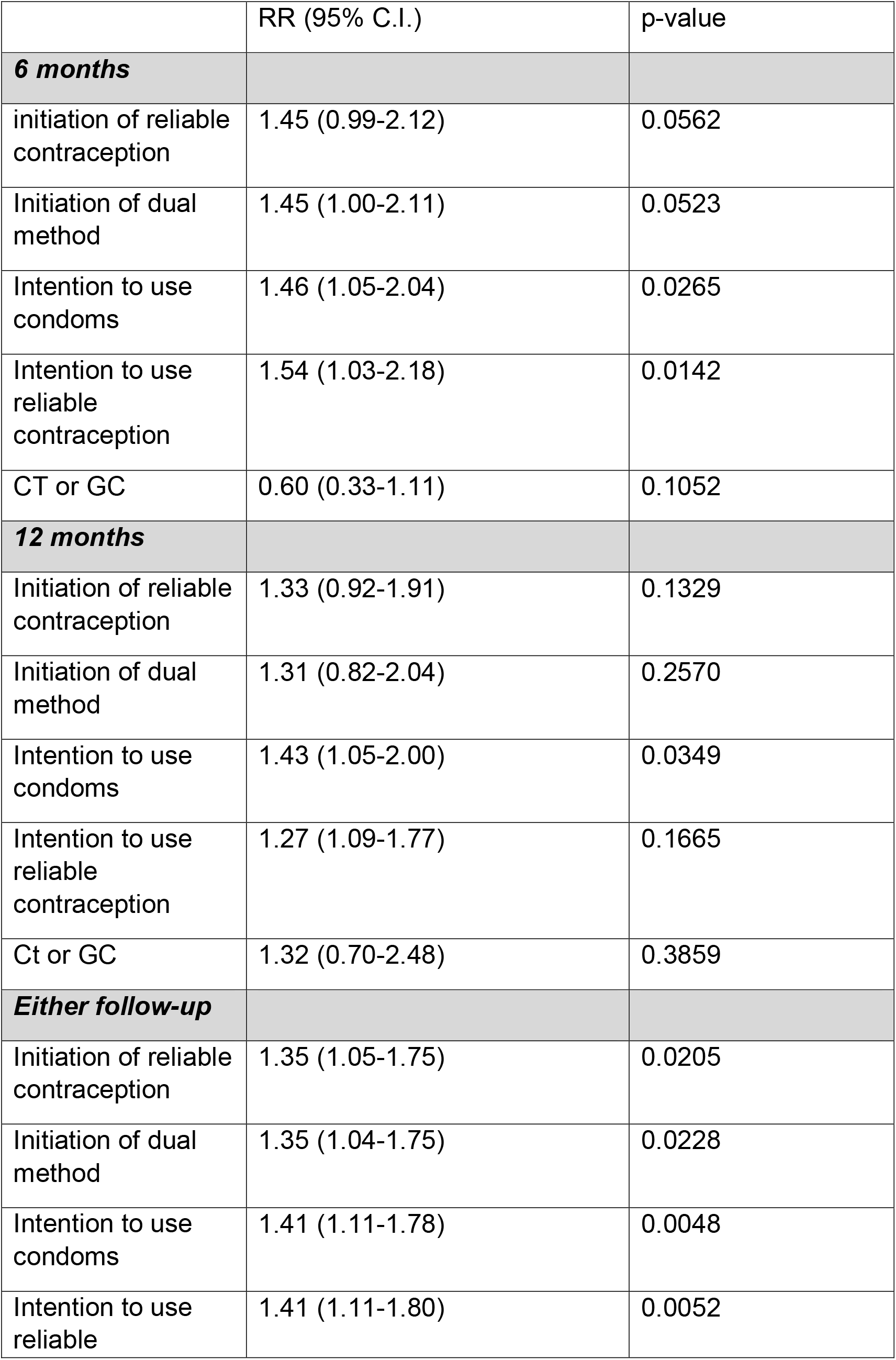
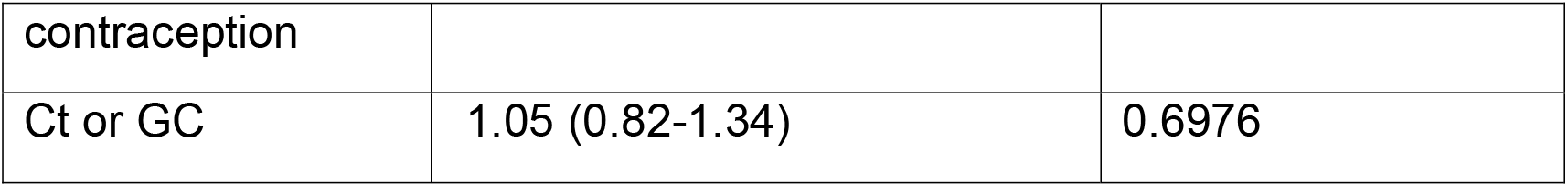
Outcomes Intent-to-treat analysis (N=637)

**Figure 2.**
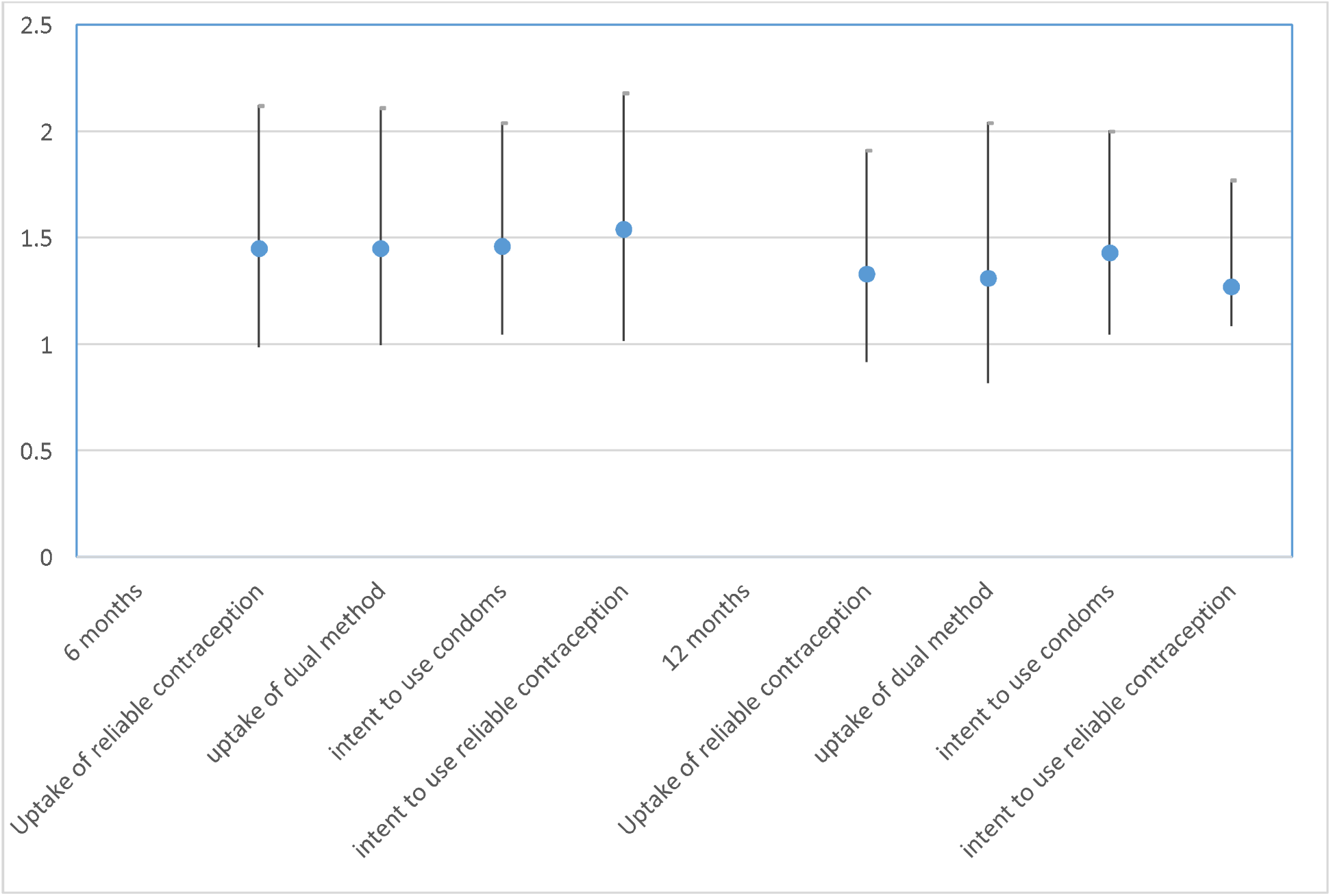
Relative Risk (intervention versus control) for primary outcomes Intent-to-treat analysis at 6 and 12 months (N=637)

### Secondary outcomes

Of the 637 participants, 569 (89.3%) were followed and were assessed for pregnancy and 27 (4.7%) unique women became pregnant. Of the 27 pregnancies, none were actively seeking to become pregnant but 2 (7.4%) said they did not mind if they became pregnant, both of whom were on the control arm. In intent-to-treat Cox Regression analysis, intervention arm was similar to control arm for becoming pregnant H.R. 0.96 (95% C.I. 0.44 – 2.03, p=0.90). In per protocol analysis, intervention trended toward a lower rate H.R. 0.52 (95% C.I. 0.02 – 1.39, p=0.17).

Of the cohort, 7.3% had an STI at 6 months and 7.2% at 12 months. Of the 72 incident STIs, 34 were among intervention women and 38 among control women. Only 4 (5.6%) of these incident STIs were self-reported. There was a non-significant trend for a lower rate of STIs among intervention compared to control at 6 months (R.R. 0.60, 95% C.I. 0.33-1.1), but this was not sustained at 12 months (6.4% vs.4.7%, R.R. 1.19, 95% C.I. 0.81-1.76). Of the 71 women who reported an STI at baseline, 37.0% reported that both they and all their partners were treated. Twenty-three women (3.6%) tested positive for an STI at a subsequent visit. There was no difference between arms for those who had an infection at a follow up visit (3.5% in the intervention vs. 3.7% in the control, p=0.57). Treatment information was not consistently collected.

### Satisfaction with program

Of the 556 (87.2% of total) women who completed their 2-month visit, 94.0% said they enjoyed/very much enjoyed the program and 98% said they would recommend it to a friend. Women in the intervention arm were more likely to have enjoyed/very much enjoyed the program compared to controls (96.7% vs.91.4%, p< 0.01) but there was no difference by arm for stating they were willing/very much willing to recommend it to friends (p=0.74).

## Discussion

The intervention had a short-term effect on initiation of reliable contraception and dual method use, as well as intention to use reliable contraception and condoms in the future. This effect attenuated somewhat over time but, in general, effects remained 20%-40% greater than the comparison arm. This effect could have an important impact in a larger population. It should be noted that these outcomes were all self-reported, but since women in both arms had a similar social desirability score and the self-reported outcomes were elicited by ACASI, it is unlikely that this reflects a social desirability bias. The efficacy of interventions for behavioral change tend to decay after time,^22, 23^ thus this intervention may need to be supplemented by a booster to retain its effect.

The biological outcomes demonstrated some reduction, but the pattern was not as clear as the behavioral outcomes. The STI rate trended lower at 6 months, but this effect was not sustained at 12 months and, in fact, trended toward a higher rate at 12 months. STIs such as Ct and GC can have high repeat infection rates, particularly if partner treatment is not performed.^24^ Since only 37% of the women reported that she and all her partners had been treated, it is possible that much of the subsequent STIs were actually reinfections by an untreated partner or improperly treated original infections. Pregnancy rates over the course of follow-up demonstrated a trend for reduction in per protocol analysis but not in intent-to-treat analysis. Pregnancy rates in this study population were on par with what would be expected for this demographic (4.7% in the intervention vs. 3.9% per U.S. rates).^25^ Importantly, the study was not powered for these secondary outcomes, so the trend we found could be meaningful. Future evaluation of the BUtiful intervention in larger populations is needed to determine the effect of the intervention on these biological outcomes.

It is possible that the intervention was effective at starting the process of behavioral change but as women acquired greater awareness of the health risk posed by unprotected sex, and adopted more health-protective intentions, they would have benefited from more intensive skills building activities. Internet interventions have demonstrated efficacy for mental health^26^ but have been less consistent for HIV prevention^27, 28^ and smoking cessation.^29^ Perhaps behavioral change that involves negotiating with others, as with sexual behavior, requires some face-to-face interaction. This face-to-face part of the intervention could be using the more traditional small group format or via synchronous interaction online (e.g., video chat). Boosters could be done by brief telephone contacts.^30^

The study population was mostly in college which may not represent the most at-risk adolescent women. However, given the high rates of STI at baseline and the high rates of reported sexual risk, they demonstrated a population in need of intervention. Also, a UTP for Black teenage women in college could derail their career plans and lead to further health disparities, so it could be argued that this is a very important group to target.

The high rate of adherence demonstrates feasibility of this type of internet intervention. It is not clear whether the financial incentive (i.e. $5 for each session completed) or the frequent text/email/phone reminders could have contributed to this high completion rate as some other electronically delivered interventions had low completion rates.^31^ Those who engage in internet interventions should build in reminders and possibly incentives to assure participation. Moreover, the use of apps and the internet has changed greatly since the time the intervention was created. New platforms such as TikTok, and increased use of behavioral change apps for fertility tracking, women’s health, etc., might increase acceptability of such an online intervention.

In sum, this internet intervention demonstrated efficacy in self-reported UTP/STI prevention behaviors and a trend for lower pregnancy rates and STIs, though the effect appears to wane with time. Replication of this study in larger samples, and possibly with supplementation of more skills building and/or booster, is needed.

## Data Availability

Data for the present study is unable to be shared

## Acknowledgements

We wish to thank the people who supported this project at our collaborating sites including: Delgado Community College, Southern University at New Orleans, Dillard University, Xavier University, and the Tulane Drop In Clinic. We thank also the staff and student research assistants: Taylor Johnson, Steffani Bangel, Brittany McBride, Jonjelyn Gamble, Braiden Eilers, Sarah Pallin, Alys Adamski, Breanna Hillman, Genevieve Gaudet, Nadira Abudru-Rahman, Yves-Yvette Young, Yewande Olugbade, Prema Bhattacherjee, Upama Aktaruzzaman, Emily Flanigan, and Brittani Coore. Special thanks to the SiHLE health educators (Tiffany Renfro and Nikia Braxton) for training us in SiHLE, New Orleans Video Access Center (NOVAC) for producing the videos, Alex Bengoa and the Tulane Technology team and Sushil Karampuri and eAbyas Info Solutions Pvt Ltd who helped us create the website, and graphic designer Katrina Andy. We are very grateful to the community advisory board and the subjects for their input and participation.

